# Head kinematics associated with off field head injury assessment (HIA1) events in a season of English elite-level club men’s and women’s rugby union matches

**DOI:** 10.1101/2024.10.01.24314695

**Authors:** David Allan, James Tooby, Lindsay Starling, Ross Tucker, Éanna Falvey, Danielle Salmon, James Brown, Sam Hudson, Keith Stokes, Ben Jones, Simon Kemp, Patrick O’Halloran, Matt Cross, Melanie Bussey, Gregory Tierney

**Author notes:** **Corresponding author:** Dr David Allan, Ulster University, Belfast, United Kingdom.

## Abstract

**Objectives:** To investigate head kinematic variables in elite men’s and women’s rugby union and their ability to predict player removal for an off-field (HIA1) head injury assessment.

**Methods:** Instrumented Mouthguard (iMG) data were collected for 250 men and 132 women from 1,865 and 807 player-matches, respectively, and synchronised to video-coded match footage. Head peak resultant linear acceleration (PLA), peak resultant angular acceleration (PAA) and peak change in angular velocity (dPAV) were extracted from each head acceleration event (HAE). HAEs were linked to documented HIA1 events, with ten logistical regression models for men and women, using a random subset of non-case HAEs, calculated to identify kinematic variables associated with HIA1 events. Receiver operating characteristic curves (ROC) were used to describe thresholds for HIA1 removal.

**Results:** Increases in PLA, and dPAV were significantly associated with an increasing likelihood of HIA1 removal in the men’s game, with an OR ranging from 1.05-1.12 and 1.13-1.18, respectively. The optimal values to maximise for both sensitivity and specificity for detecting an HIA1 were 1.96krad.s^-2^, 24.29g, and 14.75rad.s^-1^ for PAA, PLA, and dPAV respectively. Only one model had any significant variable associated with increasing the likelihood of a HIA1 removal in the women’s game – PAA with an OR of 8.51 (1.23-58.66). The optimal values for sensitivity and specificity for women were 2.01krad.s^-2^, 25.98g, and 15.38rad.s^-1^ for PAA, PLA, and dPAV respectively.

**Conclusion:** PLA and dPAV were predictive of men’s HIA1 events. Further HIA1 data are needed to understand the role of head kinematic variables in the women’s game. The calculated spectrum of sensitivity and specificity of iMG alerts for HIA1 removals in men and women present a starting point for further discussion about using iMGs as an additional trigger in the existing HIA process.

**Key Points:**

- Concussion is the most common injury in rugby union. Current on-field suspected concussion detection methods rely on visually identifying an athlete exhibiting concussion signs, reporting symptoms or identifying clinical features in real time or upon video review of the event.
- Increases in peak linear acceleration (PLA) and changes in Peak Angular Velocity (dPAV) were predictive of men’s Head Injury Assessment 1 (HIA1) events, and Peak Angular Acceleration (PAA) was predictive of women’s HIA1 events; however, further HIA1 data are needed to fully understand the role of head kinematic variables within the women’s game.
- The findings contributed to the evidence that informed the 2024 World Rugby policy change to include instrumented mouthguards (iMG) measurements as a trigger for the HIA1 removal process in the elite adult game.

## 1. Introduction

Consistently identifying suspected concussions on the field presents a clinical challenge for all sports.[1] Current on-field detection methods rely on visual identification supported by video review of potentially injurious events by sideline medical practitioners who identify impacts that result in some or all of a defined set of features, including observable signs, abnormalities of cognition, balance and ocular movement.[2] Where these signs are not observed, identification relies on player-reported symptoms after head impacts. In elite rugby union, when such events are observed, players are removed from play either permanently (when defined observable signs of concussion are seen) or temporarily for a 12-minute off-field screen when a concussion is possible, but the diagnosis is not apparent on-field.[2,3] This constitutes the Head Injury Assessment 1 (HIA1) phase of the Head Injury Assessment (HIA) process, which is then completed with two post-match diagnostic assessments using the full SCAT6 screen conducted within two hours (HIA2) and 36 to 48 hours (HIA3) of the match. Approximately 20% of subsequently diagnosed concussions, post-match, in elite-level men’s rugby union go unrecognised on-field, even though a targeted post-match review of match video found that two-thirds of these cases displayed signs and symptoms of concussion at the time.[3] Clearly, there is the potential to further improve the identification of suspected concussions on the field.

Head Acceleration Events (HAEs) can result from either direct head contact or indirectly (inertial) through body contact.[1,4] Studies have associated head kinematics during HAEs, including linear acceleration, angular acceleration, and angular velocity, with the risk of concussion, though diagnostic accuracy was not assessed.[1,5,6] A recent review emphasised the importance of measuring linear and rotational kinematics in field-based studies [1], with rotational motion of the head being a primary contributor to brain deformation.[1] A tool that may improve the identification of significant head impacts with the potential to cause concussion is the instrumented mouthguard (iMG). iMGs have proven to be superior for measuring head linear and rotational kinematics during on-field HAEs compared to soft tissue-mounted and headgear-mounted sensors, as they provide better coupling with the skull through the upper dentition.[7] As a result, iMGs are preferred for in vivo measurements of HAEs[7] and have been used in combination with qualitative video analysis in field-based studies involving rugby union, rugby league, and American football teams.[8–11]

In the men’s and women’s elite game, World Rugby, the governing body for rugby union globally, has recently introduced iMG devices to support the identification of significant head impacts. This is intended to expand removal criteria so that players who experience Peak Linear Acceleration (PLA) and Peak Angular Acceleration (PAA) magnitudes exceeding identified thresholds are removed from play for screening, with the aim of identifying a portion of the 20% of concussions that go unrecognised in matches. The thresholds currently in use were chosen based on practical implications for medical support and to minimise in-match disruption since excessively low PLA and PAA magnitudes would necessitate the frequent removal of players, overwhelming the HIA process and ultimately causing stakeholders, including players and coaches, to reject its use. As such, a combined PLA-PAA threshold of 75g and 4.5krad.s^-2^ was chosen for men and 65g and 4.5krad.s^-2^ for women. This is understood to be sub-optimal in the sense that it would compromise the sensitivity of the iMG to identify concussions but is a necessary compromise to avoid excessive medical and game disruption. The incorporation of iMGs into the HIA is not intended to be diagnostic in nature, nor to replace the current HIA screening process, but rather as an additional criterion to identify significant head impacts that do not cause the observable clinical signs and behaviour changes that currently prompt player removal.

While the above HIA1 changes have already been introduced, the research upon which they are based has not been presented. The aim of this study is to investigate head kinematic variables in elite men’s and women’s rugby union and to explore their ability to predict if a player may be required to undergo an HIA1 assessment. The sensitivity and specificity of iMG measurements in predicting HIA1 removals were also explored.

## 2. Methods

### 2.1. Study Design and Participants

A prospective observational cohort study was undertaken. All players eligible to feature in the first-team squad for elite-level Premiership (men) and Premier 15s (women) clubs during the 2022/23 season were approached to participate. This represents England’s highest level of men’s and women’s club rugby players. Of the players approached, 544 men and 255 women consented to participate and were provided with an iMG. Of these, 250 men and 132 women provided HAE data for this study.

Data was collected from domestic league, cup and European cup competitions in men (n=1,865 player-matches) and domestic league and cup competitions in women (n=807 player-matches). Each participant provided written consent, and ethical approval for the study was given by the University’s Research Ethics Committee, Ulster University (#REC-21-0061). Participants underwent 3D dental scans and received a custom-fit iMG (Prevent Biometrics, Minneapolis, MN). The iMGs have an accelerometer and gyroscope sampling at 3200Hz and a measurement range of ±200g and ±35rad.s^-1^, respectively. An infrared proximity sensor was embedded in the iMG to assess the coupling of the iMG to the upper dentition during HAEs. The validity of the Prevent Biometrics (Prevent) iMG has been demonstrated in previous studies both on-field and within laboratory settings.[12–15] The measurements of PLA and PAA have been shown to have a concordance correlation coefficient of between 0.97 and 0.98 for PLA and between 0.91 and 0.98 for PAA when compared to reference head form measurements.[13,15] The peak kinematic validity measurements are generated within laboratory settings and may not represent the iMGs’ on-field performance.

An HAE was identified when a linear acceleration trigger threshold of 8g measured at the mouthguard was exceeded upon a single axis of the iMG accelerometer.[11] The 8g threshold was selected by Prevent to limit the number of HAE cases collected from voluntary head accelerations caused by running, jumping, and cutting, which occur at or below this level of acceleration.[11] HAE kinematics were captured 10ms pre-trigger and 40ms post-trigger. For reporting HAEs, the kinematic signals are transformed to the head centre of gravity (CG), in line with SAE J211 reporting standards. [16]

As HAEs from contact events have been recorded below 10g within Rugby Union[9,15], a threshold of 400rad.s^-2^ and 5g at the head CG was utilised to capture and include HAEs from contact events only.[11] A trigger threshold of 8g can record HAEs below 8g at the head CG. The thresholds of 400rad.s^-2^ and 5g have been shown to have a Positive Predictive Value (PPV) of 0.99 (95% CI 0.97–1.00) for identifying HAEs that come directly from contact events.[11]

The resultant PLA at the head CG, resultant PAA, and the change in the head’s peak angular velocity (dPAV) were extracted from each HAE.[9] Peak change in angular velocity was calculated by zeroing the three component waveforms to the onset of post-trigger data (i.e. the moment the iMG was triggered) with the peak resultant value of dPAV calculated from the zeroed components.[9] PAA was calculated by differentiating the raw angular velocity time trace using a five-point stencil method.[18] The level of noise/artefact in the raw kinematic signal is classified by Prevent, using a proprietary algorithm, to determine whether each HAE contains minimal noise (class 0), moderate noise (class 1) or severe noise (class 2), see Limitations. A total of 47741 HAE were collected, of which 43697 (91.5%) were class 0, 3254 (6.8%) were class 1, and 790 (1.7%) were class 2. The filtering process for the Prevent iMG has previously been described in detail. In brief, a fourth-order (2x2 pole) zero phase, low-pass Butterworth filter with a cut-off frequency (−6 dB) [17] of 200, 100 and 50 Hz for class 0, 1 and 2 HAE respectively, similar to previous studies.[9,15,18,19]

### 2.2. iMG and HIA Event Identification

Removals from play for HIA1 assessments were exported from the World Rugby SCRM database.[3] The SCRM App records all clinical assessments and data related to the HIA protocol globally in a secure database. There are in-built data validation checks within the SCRM App to improve data accuracy, and World Rugby employs an independent researcher to perform weekly quality control of the data stored by SCRM to ensure the data are sufficiently accurate to use for research.

To identify the HAE event that led to a player’s removal for a HIA1 assessment, match footage and event data were obtained from StatsPerform (Chicago, Illinois, United States). Match data includes player information related to contact events, such as tackles, carries, rucks, etc., and other information, such as substitution timings. For players that were removed for a HIA1 assessment, the time they were removed from the field was used to synchronise the iMG HAE timestamps to that of the contact events. The contact events leading up to the player’s removal were then watched to identify the HAE impact responsible for HIA1 removal using the rationale that either the final HAE before the removal of the player or the most significant HAE in the moments before player removal was responsible for the HIA1. If the head impact was not clearly visible from video footage, the HIA1 case was excluded, and the potential HAE impacts which led to the player’s removal were removed from the analysis (n=4).

### 2.3. Statistical Analysis

Binary logistical regression and odd ratios (OR) with 95% CI were calculated to identify iMG kinematics associated with HIA1 events compared to non-cases (i.e. HAEs that did not lead to a HIA1). Non-case HAEs were not video verified; however, as stated above, as a 400rad.s^-2^ and 5g threshold were used, the number of false positive events, i.e. events that did not originate from contact, is expected to be very low (PPV>0.99). [11] Due to the non-independent nature of the non-cases, 10 sets of random samples (1 random non-case event per player) were taken no non-case event was included more than once across the 10 random samples. This also limited the number of observations to limit and prevent oversampling of the non-cases in relation to the HIA1s. In cases where a player was linked to multiple HIA1 events, to maintain the independence of the predictor variables and to take a conservative approach, the minimum recorded values for PLA, PAA, and dPAV values were input into the model.

A logistic regression model was run separately for all ten models for both men and women, and each model contained three independent variables: PLA, PAA, and dPAV. The multicollinearity of the independent variables (PLA, PAA, dPAV) was formally assessed.[20] No independent variables were found to be highly intercorrelated (Collinearity Tolerance all greater than 0.1, Variance Inflation Factor (VIF) less that 10).[20] The addition of interaction effects to the models was also investigated, but no evidence was found to support their inclusion and, therefore, not reported. Receiver Operator Characteristic curves (ROC) were calculated for the independent variables with optimal thresholds for player removal calculated. Optimal threshold values were determined using the Youden Index which maximises both the sensitivity and specificity of the independent variables. The non-case impacts used for the 10 random models were combined to form one singular dataset for both men and women separately, as ROC analysis does not suffer from the sparse occurrence of HIA1 cases.

Peak head kinematics linked to HIA1 events were compared between men and women using linear mixed-effects models.[9,10] Fixed effects included the categorical variable sex. The normality of the data was assessed, and a normal distribution was found through visual inspection using histograms, Shapiro-Wilk test, and Q-Q plots for the residuals. Linear mixed-effects models were used to evaluate nested data in clusters of individual players. PLA, PAA and dPAV were compared between men and women. The player subject factor was included as a random intercept for the model to account for variability. Significance was determined by comparing the p-values with an alpha level of p<0.05. Statistical analyses were conducted using commercially available software (IBM^®^ SPSS®v.29, STATA).

## 3. Results

Over the included competitions, 38,842 HAEs were collected in the men’s game, with 8,899 in the women’s game. Figure 1 shows the relationship between PLA vs PAA and dPAV vs PAA of all HAEs and identified HIA1 cases in men (left) and women (right). A total of 30 and 8 HIA1 assessments from 27 and 8 players wearing iMG were identified in the men’s and women’s cohorts, respectively.

**Figure 1.**
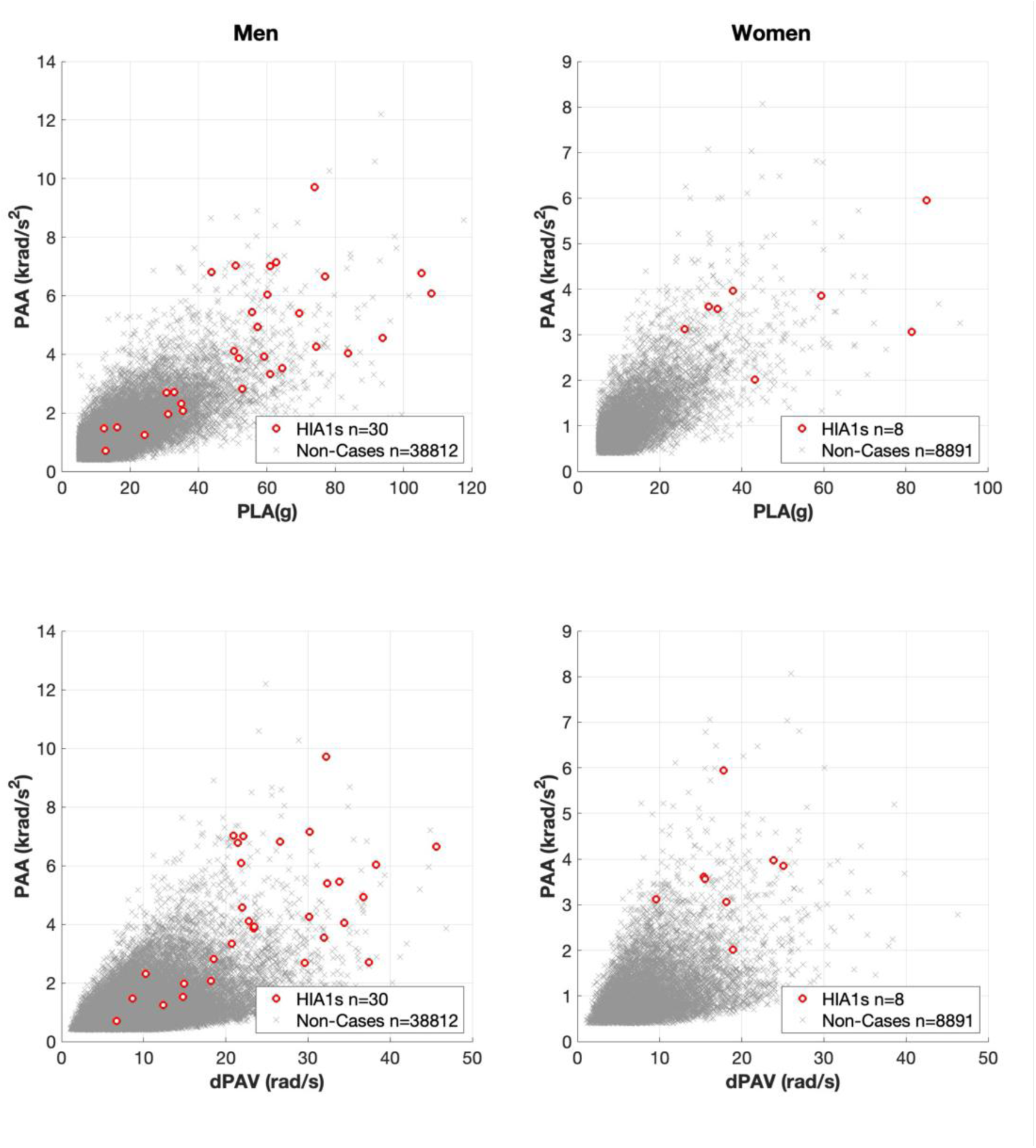
Scatter plot illustrating the kinematics values for HIA1 events relative to non-cases for men and women.

In the men’s game, the logistic regression analysis indicates that increases in PLA and dPAV are the strongest predictors of whether a player would be removed for an HIA1 assessment. Within seven of the ten models run, PLA was found to be a significant predictor (Table 1). dPAV produced significant findings in eight out of the ten models. In no model was PAA found to be significant. Six models had both PLA and dPAV as significant predictors. The Area Under Curve (AUC) ranged from 0.93-0.95 (Table 1).

**Table 1:** Results of the 10 random sample logistic regression models for the men’s game. Peak Angular Acceleration (PAA) is measured in krad.s^-2^, Peak Linear Acceleration (PLA) is measured in g, and change in Peak Angular Velocity (dPAV) is measured in rad.s^-1^. AUC represents the area under the logistic regression curve and is a measure of model accuracy.

|  | Constant | PAA | PLA | dPAV | AUC |
| --- | --- | --- | --- | --- | --- |
| Model 1 | 0.002* | 1.30 (0.67-2.52) | 1.05 (1.00-1.11) | 1.16 (1.04-1.29)* | 0.93 (0.87-0.99) |
| Model 2 | 0.002* | 1.15 (0.54-2.43) | 1.06 (1.00-1.13) | 1.18 (1.06-1.31)* | 0.94 (0.89-0.99) |
| Model 3 | 0.001* | 1.88 (0.64-5.57) | 1.07 (0.99-1.16) | 1.10 (0.99-1.22) | 0.94 (0.89-1.00) |
| Model 4 | 0.002* | 1.16 (0.57-2.36) | 1.08 (1.01-1.16)* | 1.13 (1.04-1.23)* | 0.94 (0.89-1.00) |
| Model 5 | 0.002* | 1.21 (0.56-2.62) | 1.08 (1.01-1.15)* | 1.13 (1.01-1.27)* | 0.95 (0.89-1.00) |
| Model 6 | 0.001* | 0.87 (0.41-1.85) | 1.11 (1.03-1.19)* | 1.17 (1.05-1.31)* | 0.95 (0.89-1.00) |
| Model 7 | 0.001* | 1.64 (0.67-4.01) | 1.07 (1.00-1.14)* | 1.13 (1.01-1.27)* | 0.94 (0.89-0.99) |
| Model 8 | 0.002* | 1.11 (0.60-2.05) | 1.05 (1.00-1.11)* | 1.16 (1.07-1.27)* | 0.94 (0.89-0.99) |
| Model 9 | 0.002* | 1.27 (0.55-2.92) | 1.09 (1.00-1.18)* | 1.08 (0.97-1.20) | 0.95 (0.90-1.00) |
| Model 10 | 0.002* | 0.67 (0.34-1.32) | 1.12 (1.04-1.20)* | 1.15 (1.04-1.28)* | 0.95 (0.90-1.00) |

*Significant predictors ( $p > 0.05$ ) are denoted with a \**

The results of the ROC analysis for the men’s game indicated that the PAA threshold value that optimised for both sensitivity and specificity was 1.96krad.s^-2^ (sensitivity 85.2% (66.3%-95.8%), specificity 89.2% (87.8%-90.5%)). For PLA, the highest sensitivity (88.9%, 70.8%-97.6%) and specificity (88.7%, 87.2%-90.0%) were found at a threshold of 24.3g. The dPAV threshold to maximise sensitivity and specificity was 14.75rad.s^-1^ (sensitivity, 85.2% (66.3%-95.8%), specificity 88.6% (87.1%-89.9%)). Figure 2 shows the relationship between sensitivity and specificity at a range of PLA and PAA values for men and women. As PLA increases, there is an expected reduction in sensitivity, with values declining to 11.1% at a PLA of 75g while specificity increases to 100%. For a PAA of 4.5krad.s^-2^, the sensitivity and specificity are 40.7% and 99.5%. Supplementary Figures 1 and 2 show the ROC analysis in addition to sensitivity and specificity of the predictor variables for the 10 individual models.

**Figure 2:**
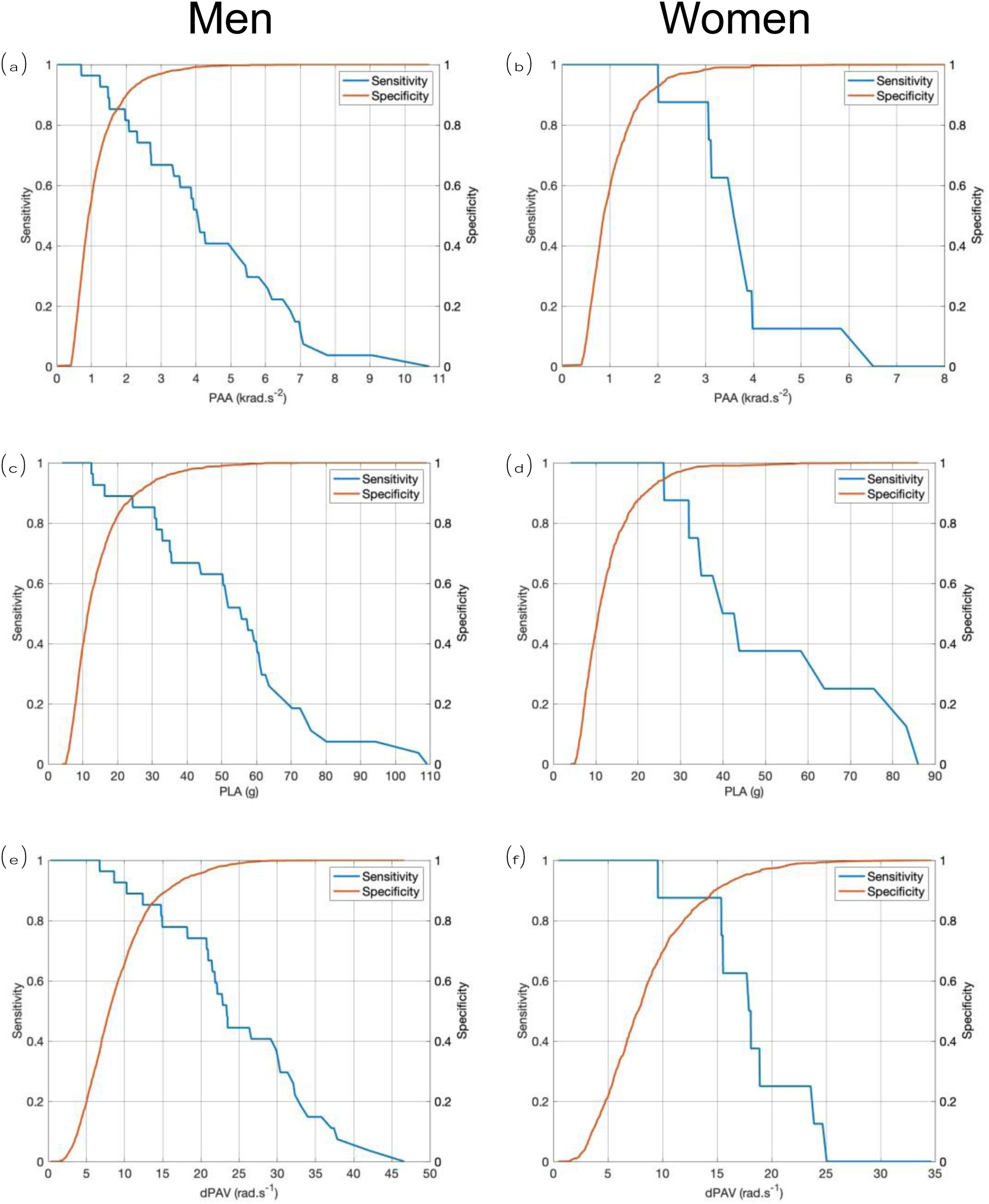
Sensitivity and Specificity based on the Receiver Operating Characteristic curve (ROC) analysis for men and women for the kinematic variables of Peak Angular Acceleration (PAA) (a&b), Peak Linear Acceleration (PLA) (c&d), and change in Peak Angular Velocity (dPAV) (e&f).

In the women’s game, eight of the ten logistic regression analyses reached a final solution. The results show a significant association for PAA and HIA1 removal within only one model (Table 2). No other models contained significant predictors. Supplementary Figure 3 shows the sensitivity and specificity of the predictor variables for the 8 individual models that reached a final solution.

**Table 2:** Results of the 8 random sample logistic regression models that reached a final solution for the women’s game. Peak Angular Acceleration (PAA) is measured in krad.s^-2^, Peak Linear Acceleration (PLA) is measured in g, and change in Peak Angular Velocity (dPAV) is measured in rad.s^-1^. AUC represents the area under the logistic regression curve and is a measure of model accuracy.

|  | Constant | PAA | PLA | dPAV | AUC |
| --- | --- | --- | --- | --- | --- |
| Model 1 | 0.000* | 2.38 (0.76-7.45) | 1.03 (0.96-1.10) | 1.20 (0.91-1.60) | 0.97 (0.94-1.00) |
| Model 2 | 0.000 | 33.04 (0.31-3509.62) | 1.33 (0.92-1.92) | 1.07 (0.66-1.75) | 1.00 (0.99-1.00) |
| Model 3 | 0.000 | 129.43 (0.20-84130.34) | 1.84 (0.85-3.98) | 0.56 (0.24-1.31) | 1.00 (0.99-1.00) |
| Model 4 | 0.000 | 13.13 (0.75-230.06) | 1.39 (0.84-2.29) | 0.73 (0.39-1.36) | 1.00 (0.99-1.00) |
| Model 5 | 0.002* | 1.65 (0.74-3.72) | 1.12 (1.00-1.26) | 0.97 (0.75-1.25) | 0.98 (0.96-1.00) |
| Model 6 | 0.000 | 81.88 (0.11-58555.78) | 1.08 (0.89-1.31) | 1.52 (0.64-3.60) | 1.00 (0.99-1.00) |
| Model 7 | 0.001* | 2.71 (0.96-7.60) | 1.08 (0.97-1.20) | 1.04 (0.80-1.36) | 0.98 (0.96-1.00) |
| Model 8 | 0.000* | 8.51 (1.23-58.66)* | 1.03 (0.93-1.14) | 1.24 (0.94-1.65) | 0.99 (0.98-1.00) |

*Significant predictors ( $p > 0.05$ ) are denoted with a \**

The results of the ROC analysis for the women’s game indicated that the PAA threshold value that optimised for both sensitivity and specificity was 2.0krad.s^-2^ (sensitivity, 100% (68.8%-100%), specificity 92.8% (91.0%-94.3%)). The threshold value for PLA was 26.0g (sensitivity, 100% (68.8%-100%), specificity (94.5% (92.9%-95.8%)) while the optimal threshold for dPAV occurred at 15.4rad.s^-1^ (sensitivity, 87.5% (47.3%-99.7%), specificity 91.3% (89.5%-93.0%)) (Figure 2). At the PLA and PAA thresholds currently in use (65g and 4.5krad.s^-2^), sensitivity decreased to 25% and 12.5%, while specificity increased to 99.8% and 99.7%, respectively.

No significant differences in PLA (p=0.61), PAA (p=0.40), and dPAV (p=0.06) were found between men’s and women’s HIA1 events. Mean and standard deviation (SD) PLA, PAA and dPAV for HIA1 events were 54.9g (SD=24.9), 4.3krad.s^-2^ (SD=2.2) and 24.8rad.s^-1^ (SD = 9.6) for men respectively, and 49.8g (SD=22.8), 3.6krad.s^-2^ (SD=1.1) and 18.0rad.s^-1^ (SD=4.9) for women, respectively. Mean PLA, PAA and dPAV for non-case HAE events were 14.4g (SD=9.2), 1.1krad.s^-2^ (SD=0.7) and 9.0rad.s^-1^ (SD = 4.9) for men, respectively, and 13.3g (SD=8.0), 1.1krad.s^-2^ (SD=0.7) and 9.0rad.s^-1^(SD=4.8) for women, respectively.

## 4. Discussion

The present study finds associations between head kinematics, specifically PLA and dPAV in men and PAA in women, and HIA1 removals during match play in professional rugby union from a dataset of over 47,000 HAEs and 38 HIA1s. The association between these head kinematic variables and HIA1 removals invites the potential use of iMG as a means to identify significant head impacts that may warrant clinical assessment as part of the sport’s HIA process and is the data set that contributed to World Rugby’s addition of iMG into its HIA process in 2024.

We find that PLA and dPAV predict men’s HIA1 events in nine of ten random sample logistic regression models, with further HIA1 data needed to understand the role of head kinematic variables within the women’s game. Adding the iMG to the removal-from-play criteria reduces the reliance on player self-report and visual observations from side-line medical practitioners and/or video reviewers to visually identify a suspected concussive impact on the field. The addition of iMG may thus reduce the proportion of concussions that are not identified at the time of the inciting event.[3]

The results of the ten logistic regression models for the men show significant associations between the HAE kinematics of PLA and dPAV and the odds of a player being removed for an HIA1 assessment. Of these models, nine were found to have significant predictors; one model had only PLA being predictive, two models had only dPAV being predictive, and six models with PLA and dPAV being predictive (Table 1). The OR for the significant PLA predictors ranges from 1.05 to 1.12, with the OR of the significant dPAV predictors ranging from 1.13 to 1.18. The OR imply that for every 1g increase in PLA increases the odds of a HIA1 removal increase by between 5% and 12%; for dPAV, the OR implies that for every 1rad.s^-1^ increase the odds of an HIA1 removal increase by between 13% and 18%.

The relationship between clinical outcomes of head injury and the magnitude of head accelerations has been suggested previously.[1,5,6] Data from helmet sensor field-based studies have indicated that rotational acceleration, in particular, is associated with concussion [21,22]. Other studies using helmet sensors found rotational acceleration is a significantly worse predictor of concussion than linear acceleration.[23] For the men PAA was not significantly associated with HIA1 removal in any of the models in the current study.[23] Further research is needed to understand the interaction of linear and rotational kinematics and their influence on rugby union concussions.

The results of the ten logistic regression models for the women, of which only eight reached significance, only showed a significant association with PAA within one model with an OR of 8.51 (1.23-58.66). The lack of agreement between the two cohorts may be due to a relatively small sample size of women, specifically the number of HIA1 cases available for analysis. The limited sample size of the HIA1 cases has most likely led to the women’s cohort being statistically underpowered for this analysis, and future research with larger cohorts, specifically women, is required to explore head kinematic relationships further.

The purpose of this logistic regression analysis is not to derive any form of diagnostic tests, nor is it to propose thresholds that should be used to identify HIA1 removals. Rather, these data represent the first stage of understanding what parameters may be predictive of a player requiring to undergo an HIA1 assessment. As World Rugby collects more HAE data, particularly HAEs associated with HIA1, it is envisaged that these analyses be revisited with additional parameters to identify potential further predictive parameters. The use of iMGs is not a replacement for the current HIA process but an additional tool to aid in identifying players who should be removed from play at the time of the head impact and then screened within the sport’s existing diagnostic processes.

The ROC analysis undertaken for the independent variables of PAA, PLA, and dPAV investigates how the sensitivity and specificity of the independent variables relates to the potential diagnostic performance of head kinematics in identifying HIA1 removals. It must be emphasised that this is assessed not from a diagnostic perspective but rather for removal of play as a HIA1 case. That is, our analysis is concerned solely with whether a player left the field to enter the HIA pathway at the time of a head impact.

Our finding in this regard is that the threshold values for PLA and PAA that optimised sensitivity and specificity (PLA 24.3g in men and 26.0g in women, and PAA 1.96krad.s^-2^ in men and 2.01krad.s^-2^ in women) produce good overall diagnostic accuracy. For the men, at these thresholds, a sensitivity of 88.9% and a specificity of 88.7% for PLA, and a sensitivity of 85.2% and a specificity of 89.2% for PAA were found. For the women, a sensitivity of 100% and a specificity of 94.5% for PLA, and a sensitivity of 100% and a specificity of 92.8% for PAA were found (Figure 2).

However, whether these thresholds are practical and feasible for use within the game context is a separate consideration. Using iMGs as part of the removal from play criteria cannot be concerned solely with the theoretical optimisation of sensitivity and specificity. It must also recognise that removing players from the field of play is disruptive, and so, should be reduced to an absolute tolerable minimum. Failure to do so would result in numerous instances where iMG alerts cause players to be removed for HIA1s, later found to be false positive cases. A high rate of false positive cases would create the risks of directly overwhelming medical support staff and infrastructure during matches and disrupting the match to such an extent that the premise of iMG use is rejected by coaches and players.

Based on previously published data, the volume of HAEs that occur during a match at the thresholds identified as optimising diagnostic performance would indeed cause this excessive abundance of HIA1 removals. Tooby et al reported approximately 6 HAEs over 20g per player hour and 2.8 HAEs per player hour over 2krad.s^-2^ for men and 2.4 HAEs per player hour over 20g and 1.5 HAEs per player hour over 2krad.s^-2^ for women. If every player wore the iMG, it can be predicted that over 100 iMG alerts would regularly occur in the men’s elite game, for example. To reduce the number of iMG alerts that would need assessing by medical staff and the subsequent potential overall disruption to the game, a compromise is needed whereby the thresholds are adjusted to minimize false positive occurrences (i.e. maximize specificity). This will reduce sensitivity, meaning that potentially injurious head impacts are missed by the IMG. However, many of these events should be detected by match day officials, doctors, and medical personnel, with the result that an increase in specificity does not have a large effect on missed cases, but is necessary for adoption into the sport’s HIA protocol

Figure 2 shows this trade-off as the PLA and PAA thresholds are increased to the levels that World Rugby has chosen for implementation in the elite game. The PLA threshold of 75g has a sensitivity of 11.1% and specificity of 100%, and the PAA threshold of 4.5krad.s^-2^ has a sensitivity of 40.7% and specificity of 99.5%. For women, the PLA threshold of 65g has a sensitivity of 25% and specificity of 99.8%, and the PAA threshold of 4.5krad.s^-2^ has a sensitivity of 12.5% and specificity of 99.7%. Further evaluation of the performance of the iMG in this regard, as well as the clinical strategy for using iMG by the sport, is beyond the scope of this manuscript.

Further, our analysis assesses how head kinematics are associated with removal from play in the HIA1 phase of the HIA process. The association between head kinematics and removal from play is the most relevant clinical outcome at present since it is for HIA1 removal from play that the use of iMGs is being added. That is, iMG is additive to the current criteria for player removal. iMG is not intended to diagnose concussion, though future research may explore how PLA, PAA, dPAV and other metrics are associated with clinical outcomes. Therefore, continuing to build and develop the model as more HIA1 kinematic data becomes available has the potential to develop greater predictive capabilities.

High PLA, PAA and dPAV measures were observed in non-clinical cases (Figure 1) but without the real-time observation of clinical signs, symptoms or behaviour changes, which may be genuinely absent, or it may be that the player has continued to play without disclosing or exhibiting any consequences of the HAE. We are unaware whether these HAEs resulted in clinical presentation of signs and symptoms post-match. However, this should be a focus of future work. Whilst the use of HIA player removal thresholds is still in its infancy, this study does provide some initial starting data regarding potential predictive head kinematics and thresholds to better support the removal of players from the field of play who, based on their HAE kinematics may be at risk of a concussion.

### 4.1. Limitations

Player compliance in wearing the iMGs posed a significant challenge over the duration of this study. Out of the 530 and 232 male and female participants who consented to participate and were provided with an iMG, data was collected from 255 and 133 individual players, respectively. The challenges with player compliance may introduce a bias because those players who chose to wear the iMG may adopt different behaviours compared to players who did not wear the device. The nature of this bias is uncertain.

The current study may not fully represent the various playing styles and conditions found in all levels of rugby worldwide. HIA kinematics may differ in other rugby cohorts, particularly in the adult and age-group amateur and community games.

The present study used peak resultant head kinematics but did not consider the directionality and temporal aspects (e.g., pulse duration) of the kinematic signals obtained from the iMG. Temporal and directional factors are likely crucial in understanding injury risk and should be included in future work, particularly for the relationship between HAE kinematics and clinical outcomes. The kinematic signal processing was conducted by the Prevent system, similar to other commercially available iMG systems.[15] The kinematic signal processing utilised in the current study has been incorporated into validations of the Prevent iMG system [15] and is currently utilised in professional rugby. However, a common and agreed-upon signal processing approach for iMG systems, such as the HEADSport filter method, may be warranted.[17,24] A common signal processing approach is important for future inter-study comparisons within and between sports, particularly if different iMG systems have been utilised.[17]

As explained above, we have used HIA1 removal from play as the case for the present analyses, rather than concussions, because we wished to assess the potential of the iMG as additive to this phase of the process rather than the diagnostic phases. In future studies, clinical outcomes from the entire HIA process will allow the diagnostic performance of the iMG for concussions to be assessed. However, we believe that it is appropriate for this study to evaluate the HIA1 cases because the premise of the iMG mandate by World Rugby is to use the iMG only as part of the criteria identifying players who require the HIA1 screen and not for diagnosis of concussion. This approach also enables a larger sample size for evaluation, and in future, a combined approach that assesses the associations between HAE magnitude and HIA1 indicators, as well as concussion outcomes, can be explored.

## 5. Conclusion

PLA and dPAV were found to be predictive of men’s HIA1 events in nine of ten random sample logistic regression models. Due to the low number of HIA1s within the women’s cohort, further HIA1 data are needed to understand the role of head kinematic variables within the women’s game. Only one model for women had any significant predictors, with PAA being predictive; however, it had large confidence intervals. The thresholds for both men and women present a starting point for further discussion about using iMGs as an addition to the HIA process and when removing a player for further screening and assessment may be appropriate.

## 6. Policy Implications

The findings of this paper have informed the adoption of a World Rugby policy that includes iMG measurements in the HIA1 removal process in the elite game.

## Contributors

DA, LS, RT, EF, GT conceptualised the research project. All authors were involved in the design and data collection for the study. DA, JT, LS, RT, EF, GT were responsible for the analysis and interpretation of the results. DA and GT drafted the manuscript. All authors critically reviewed and edited the manuscript prior to submission.

## Data Availability

Anonymised data available upon reasonable request.

## Data sharing

Anonymised data available upon reasonable request.

## Ethical approval

This project was approved by the University’s Research Ethics Committee, University of Ulster (#REC-21-0061). The study was performed in accordance with the standards of ethics outlined in the Declaration of Helsinki.

## Consent to participate

All participants provided written consent.

## Patient and public involvement

Patients and/or the public were not involved in the design, conduct, reporting, or dissemination plans of this research.

## Acknowledgements

The authors would like to thank all staff and players at the participating clubs (Bath Rugby, Bristol Bears Rugby, Exeter Chiefs, Gloucester Rugby, Harlequins FC, Leicester Tigers, London Irish RFC, Newcastle Falcons, Northampton Saints, Sale Sharks, and Saracens, Bristol Bears Women, Darlington Mowden Park DMP Sharks RFC, Exeter Chiefs Women, Gloucester-Hartpury Women RFC, Harlequins Women, Loughborough Lightning, Sale Sharks Women, Saracens Women, Worcester Warriors Women, Wasps Women) for their time and involvement in this study. The authors would also like to thank OPTA Sports for providing the authors access to their StatsPerform platform. The Rugby Players Association were supportive, endorsed and helped promote the study. The authors want to acknowledge Prevent Biometrics for their support and cooperation during the study. Finally, the authors would like to thank Martin Kidd, an independent statistician at the University of Stellenbosch, South Africa, for providing his assistance and expertise regarding the statistical analysis undertaken for this study.

