## Supplementary Figures for "Head kinematics associated with off field head injury assessment (HIA1) events in a season of English elite-level club men’s and women’s rugby union matches"

Short Title: Head Kinematics Associated with HIA1 Removal Events In English Rugby Union

David Allan[1], James Tooby[2], Lindsay Starling[3,4], Ross Tucker[3,5], Éanna Falvey[3,6], Danielle Salmon[3], James Brown[2,5], Sam Hudson[4], Keith Stokes[4,7], Ben Jones,[2,8,9,10,11], Simon Kemp[7,12], Patrick O'Halloran[13,14], Matt Cross[2,8], Melanie Bussey[15], Gregory Tierney[1]

[1] Nanotechnology and Integrated Bioengineering Centre (NIBEC), School of Engineering, Ulster University, Belfast, United Kingdom

[2] Carnegie Applied Rugby Research (CARR) Centre, Carnegie School of Sport, Leeds Beckett University, Leeds, United Kingdom

[3] World Rugby, 8-10 Pembroke St., Dublin, Ireland

[4] UK Collaborating Centre on Injury and Illness Prevention in Sport (UKCCIIS), University of Bath, United Kingdom

[5] Institute of Sport and Exercise Medicine, Stellenbosch University, South Africa

[6] School of Medicine & Health, University College Cork, Cork, Ireland

[7] Rugby Football Union, Twickenham, United Kingdom

[8] Premiership Rugby, London, United Kingdom

[9] England Performance Unit, Rugby Football League, Manchester, United Kingdom

[10] School of Behavioural and Health Sciences, Faculty of Health Sciences, Australian Catholic University, Brisbane, QLD, Australia

[11] Division of Physiological Sciences and Health through Physical Activity, Lifestyle and Sport Research Centre, Department of Human Biology, Faculty of Health Sciences, University of Cape Town, Cape Town, South Africa

[12] London School of Hygiene and Tropical Medicine, London, United Kingdom

[13] Sport and Exercise Medicine Service, University Hospitals Birmingham, United Kingdom

[14] Marker Diagnostics UK Ltd, United Kingdom

[15] School of Physical Education Sport and Exercise Sciences, University of Otago, Dunedin, New Zealand

**Corresponding author:** Dr David Allan, Ulster University, Belfast, United Kingdom.

**Supplementary Figures**

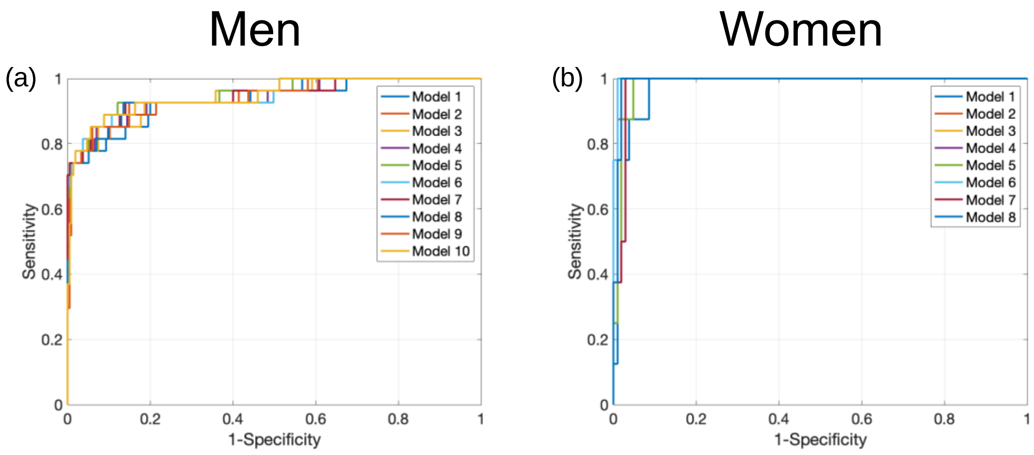


Supplementary Figure 1. Receiver Operating Characteristic (ROC) curve for the men (a) and women (b) for each of the individual logistic regression models.

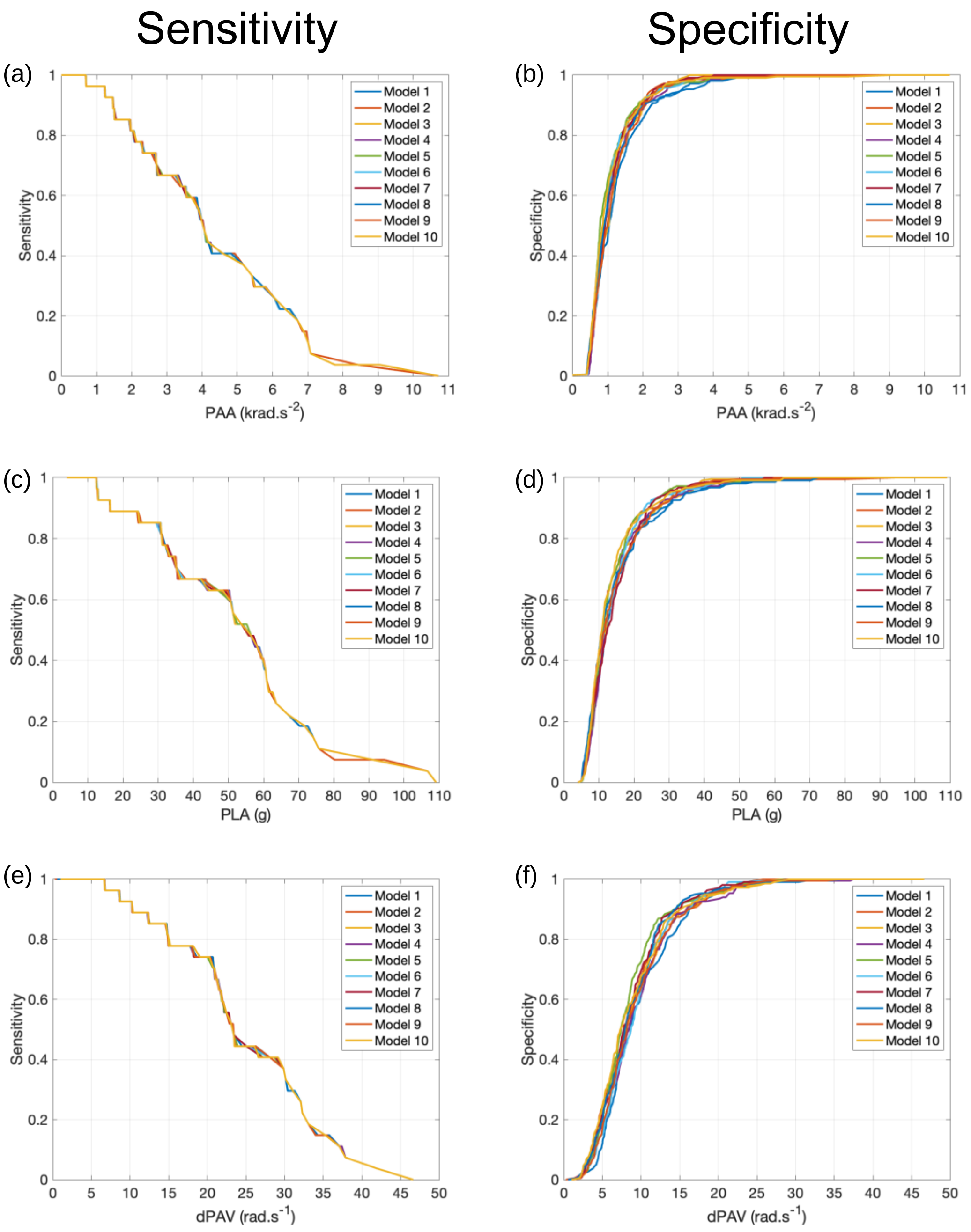


Supplementary Figure 2. Sensitivity (a,c,e) and Specificity (b,d,e) curves for the kinematic variables Peak Angular Acceleration (PAA) (a,b), Peak Linear Acceleration (PLA) (c,d), and change in Peak Angular Velocity (dPAV) (e,f) used within each logistic regression model for the men.

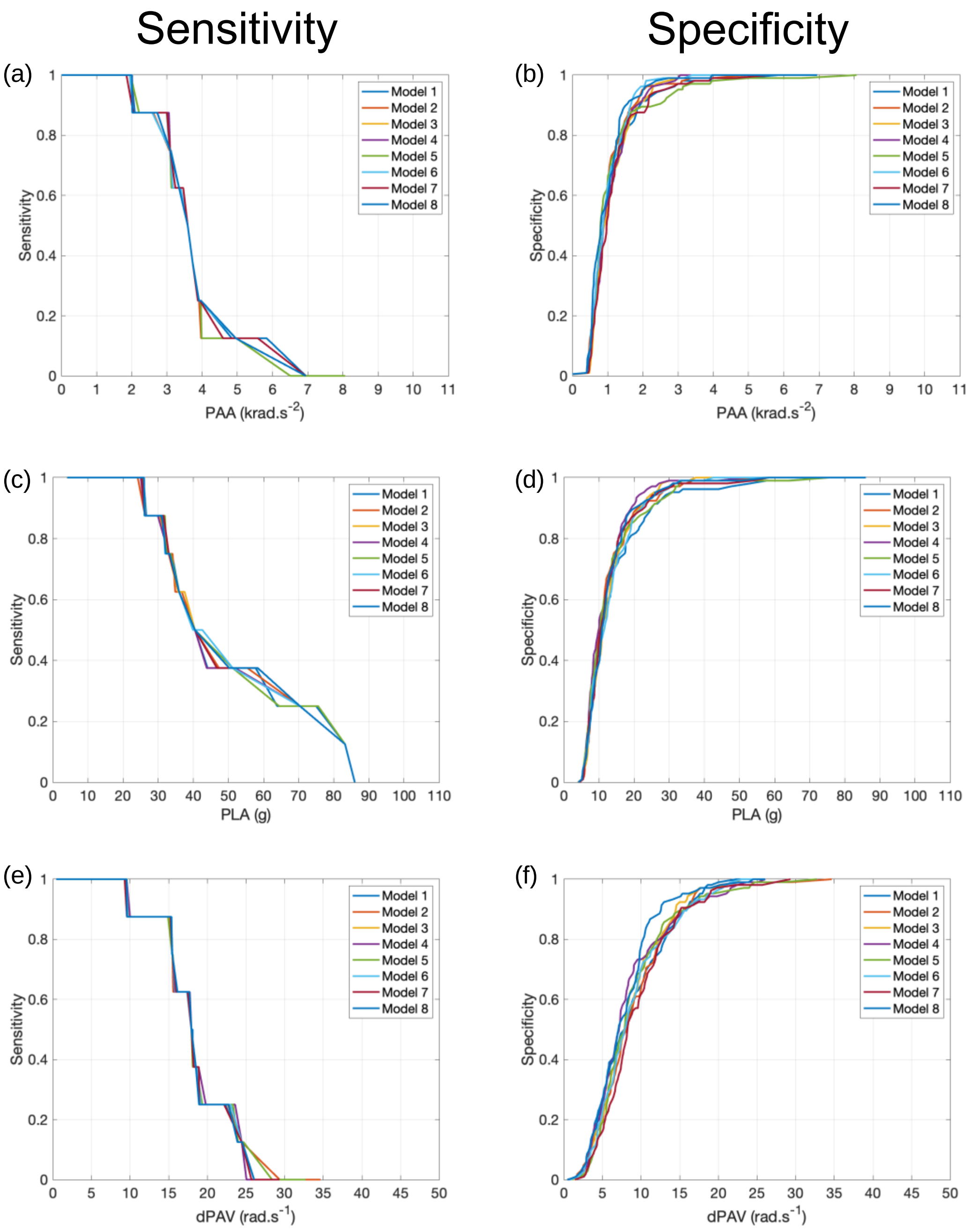


Supplementary Figure 3. Sensitivity (a,c,e) and Specificity (b,d,e) curves for the kinematic variables Peak Angular Acceleration (PAA) (a,b), Peak Linear Acceleration (PLA) (c,d), and change in Peak Angular Velocity (dPAV) (e,f) used within each logistic regression model for the men.
